# Towards AI-based Precision Rehabilitation via Contextual Model-based Reinforcement Learning

**DOI:** 10.1101/2025.01.13.24319196

**Authors:** Dongze Ye, Haipeng Luo, Carolee Winstein, Nicolas Schweighofer

## Abstract

**Background:** Stroke is a condition marked by considerable variability in lesions, recovery trajectories, and responses to therapy. Consequently, precision medicine in rehabilitation post-stroke, which aims to deliver the “right intervention, at the right time, in the right setting, for the right person,” is essential for optimizing stroke recovery. Although Artificial Intelligence (AI) has been effectively utilized in other medical fields, such as cancer and sepsis treatments, no current AI system is designed to tailor and continuously refine rehabilitation plans post-stroke.

**Methods:** We propose a novel AI-based decision-support system for precision rehabilitation that uses Reinforcement Learning (RL) to personalize the treatment plan. Specifically, our system iteratively adjusts the sequential treatment plan—timing, dosage, and intensity— to maximize long-term outcomes based on a patient model that includes covariate data (the context). The system collaborates with clinicians and people with stroke to customize the recommended plan based on clinical judgment, constraints, and preferences. To achieve this goal, we propose *a Contextual Markov Decision Process (CMDP)* framework and a novel hierarchical Bayesian model-based RL algorithm, named *Posterior Sampling for Contextual RL* (PSCRL), that discovers and continuously adjusts near-optimal sequential treatments by efficiently balancing exploitation and exploration while respecting constraints and preferences.

**Results:** We implemented and validated our precision rehabilitation system in simulations with a sequence of 100 diverse, synthetic patients. Simulation results showed the system ability to continuously learn from both upcoming data from the current patient and a database of past patients via Bayesian hierarchical modeling. Specifically, the algorithm’s sequential treatment recommendations became increasingly more effective in improving functional gains for each patient over time and across the synthetic patient population.

**Conclusions:** Our novel AI-based precision rehabilitation system based on contextual model-based reinforcement learning has the potential to play a key role in novel learning health systems in rehabilitation.

## Background

Despite extensive rehabilitation research, including multiple multi-site randomized clinical trials, about 40% of the 800,000 people who suffer a stroke in the US each year show limited recovery of upper extremity (UE) function, restricting daily activities and the quality of life(1-3). The challenge of determining optimal rehabilitation based on an individual’s clinical profile is one of the most challenging questions in stroke rehabilitation(4). Precision rehabilitation, defined as the “right intervention, at the right time, in the right setting, for the right person,” (5, 6) has been proposed as a solution to improve UE function. However, as we review below, delivering true precision rehabilitation, that is, determining the optimal sequential treatment plan that maximizes long-term outcomes for each patient, is difficult even for experienced clinicians because of the huge number of potential plans given the multiple scheduling factors that modulate recovery, the high between-patient variability, and the multiple scheduling constraints.

Here, we therefore propose a collaborative Artificial Intelligence (AI) precision rehabilitation system for stroke survivors with upper extremity (UE) deficits that uses model-based Reinforcement Learning (RL). Such model-based RL systems are being deployed in precision medicine, for instance, for cancer and sepsis treatments (7-10). However, no AI system exists to personalize and continuously refine rehabilitation treatment plans post-stroke. The collaborative AI system iteratively adjusts and recommends the plan—timing, dosage, intensity—of UE task practice based on the patient’s profile to enhance long-term outcomes; clinicians and patients can customize the recommended plan based on clinical judgment, constraints, and preferences. The system’s overall output is a personalized long-term (e.g. 6-month) treatment plan updated at each clinician-patient session. The system is self-improving both at the patient and population level: as the model is updated from a growing database, the treatment recommendations become increasingly effective in improving functional gains for each patient over the treatment horizon (i.e., duration) and across the patient population.

### Organization

This paper is organized as follows: First, we review prior research demonstrating that optimizing the longitudinal rehabilitation plan is essential for achieving the best outcomes post-stroke. However, given the multiple scheduling factors that modulate recovery, the high between-patient variability, and the multiple scheduling constraints, we argue that clinician expertise alone is insufficient to determine optimal rehabilitation plans for each patient. Second, we propose to address these challenges with a new framework for collaborative AI precision rehabilitation based on contextual model-based RL, where the context is the set of individual factors that modulate recovery. Third, given the uncertainty in the patient model, context, and scheduling constraints, we propose a novel algorithm for precision rehabilitation, *Posterior Sampling for Contextual Reinforcement Learning* (PSCRL), which generates increasingly better personalized treatment plans as the database grows. Fourth, to illustrate the functioning of the algorithm, we propose a realistic scenario of precision rehabilitation for dose scheduling. Fifth, we present simulation results to illustrate how the treatment plan continuously improves via both within-patient and between-patient learning. Finally, we discuss related work, limitations, and future clinical implementation.

### The challenges of determining the treatment plan in precision rehabilitation

We limit our AI-based precision rehabilitation system to the scheduling of UE motor rehabilitation post-stroke, which is sequential and delivered over months. Such a system assumes that optimizing the individual rehabilitation plan matters. We review four types of challenges. First, stroke recovery depends on time-varying (dynamical) processes operating at different time scales modulated by treatment parameters. Second, the effects of treatment depend on multiple individual factors, which we collectively call the “context,” that need to be considered for personalizing the rehabilitation plan. Third, not all treatments are possible; instead, they are constrained by clinical constraints, logistic constraints, and personal preferences, which we collectively call “constraints.” Finally, given the sequential nature of rehabilitation over extended periods, the number of rehabilitation plans is very large. Thus, determining the plan that maximizes long-term outcomes, given the scheduling parameters, the individual differences, and the constraints, can be best achieved by an AI system in collaboration with the clinician.

#### The dynamical processes of recovery post-stroke are modulated by treatment parameters

Stroke recovery operates via multiple time-dependent processes. The initial changes in sensorimotor behavior largely result from “spontaneous recovery”, which involves the reduction in edema, ischemic penumbra, and brain re-organization(11, 12), and is the greatest in the first month but continues for up to 6 months(13, 14). Then, motor practice can further improve sensorimotor behavior via neural plasticity mechanisms(15). However, practice affects recovery in a complex manner, with the following treatment parameters influencing the effectiveness of practice post-stroke. 1) The dose of rehabilitation, whereby high doses consisting of 1000s of trials delivered over days of practice, leads to structural and stable changes in brain areas involved in recovery(15) and to greater functional improvements(16-20). 2) The intensity of practice, whereby high daily doses enhance synaptic plasticity(15) that facilitates recovery(21, 22). 3) The timing of rehabilitation is important as motor practice that is too early or too late has been associated with worse outcomes(23, 24),(12, 25), indicating a critical “window of plasticity” (11, 12, 16, 17, 23-26), which is about 30 to 90 days for UE function in humans(27). In addition, intensive initial practice increases the probability of habit formation of motor training and, thus, long-term perseverance(28). 4) The distribution of practice is often needed (29) because a lack of sustained practice results in decreased activity in relevant motor areas(15) and loss of the gains due to rehabilitation (30). In addition, distributed practice enhances long-term performance compared to massed practice in motor learning in healthy(31) and stroke(32) populations. 5) Finally, the amount of UE use in daily activities, if above a threshold, can act as “self-training” (33-35), increasing future use and function(36, 37).

This prior research, therefore, shows that to maximize long-term outcomes, the rehabilitation plan needed to optimize recovery cannot be uniform or follow some simple predefined schedule but instead needs to be carefully crafted throughout rehabilitation in terms of dosage, intensity, timing, and distribution over time to maximize long-term gains.

#### The context associated with the variability of response to treatment

Stroke is characterized by large variability in lesions, impairments, and response to recovery(38, 39). Individuals post-stroke show highly variable responses, even to the same treatment. For instance, in re-analyses of the EXCITE(24) and DOSE(20) trials data, we found that about one-fourth of participants continued to see improvements following rehabilitation, and another one-fourth lost most gains(35, 40). The following characteristics have been shown to modulate the effect of rehabilitation: 1) lacunar and non-lacunar strokes(41); 2) baseline clinical scores(29, 42); 3) the side of lesion(43); 4) the integrity of the ipsilesional corticospinal tracts(41, 44-47); 5) somatosensory deficits(48-51) and 6) deficits in the integrity of visuospatial working memory(52), which we showed modulates the effect of massed but not distributed practice in chronic stroke(32).^a^ Therefore, the treatment plan needs to be individualized to account for the individual level clinical factors that are known to modulate the effect of treatment.

#### The constraints on possible treatments plans

Because clinical and logistic constraints as well as patient preferences limit the plans that can be delivered, not all treatment plans are possible. Clinical constraints include high activity-dependent fatigability(57), reduced attention soon after stroke, limited UE function before sufficient spontaneous recovery, and other medical conditions (e.g., shoulder pain, depression). Logistic constraints include patient schedule changes (e.g., vacations), socioeconomic and interpersonal needs, and reimbursement needs. Finally, patient preferences in scheduling and task practice are important to maximize motivation and even gains in rehabilitation. For instance, providing choices of task practice has been shown to increase gains in motor learning and rehabilitation^57^ and increase engagement and adherence(58, 59). Thus, the treatment plan must consider scheduling constraints, which we classify into clinical constraints, logistical constraints, and patient preferences.

#### The current limitations in optimizing the treatment plan

The current state-of-the-art in scheduling of task-oriented motor therapy to maximize recovery is based on the clinician’s knowledge and experience. Via both formal and continuing education, clinicians learn what treatment plan “works” best for sub-types of patients. As they treat patients and observe progress, clinicians gradually build a mental model of effective treatment plans for different sub-types of patients. However, as reviewed above, the nuances of the effect of scheduling parameters on recovery at different times post-stroke, the considerable between-patient variability, and all possible constraints make it hard to predict how patients will respond to different treatment plans and then nearly impossible to select the most effective treatment plans. Indeed, the number of potential treatment plans is huge. For instance, even the seemingly simple task of deciding whether to treat or not each week for 6 months results in ∼67 million treatment plans. Thus, it is currently not feasible to determine the treatment plan that will maximize recovery for each patient. Here, we propose that a collaborative AI-based system can help the clinician-patient team determine effective treatment plans.

## Methods

### Addressing the challenges of precision rehabilitation with Contextual Model-based Reinforcement Learning

#### Precision rehabilitation as an RL problem in a Markov Decision Process (MDP)

Although ad-hoc rehabilitation plans can be determined, we propose a structured theoretical framework for optimizing rehabilitation treatments using Reinforcement Learning (RL). Precision rehabilitation can be viewed as a decision-making problem in which the clinician-AI team interact with a person with stroke sequentially with the goal of determining treatment plans that maximize sensorimotor outcomes. The quality of treatments can be measured by longitudinal rehabilitation outcomes, which are stochastic and partly controllable, i.e., the outcomes are influenced but not fully determined by the treatment.

An appropriate and well-studied framework for such a decision-making problem is the Markov Decision Process (MDP) in the RL literature(60, 61). An MDP consists of four primary components (𝒮, 𝒜, 𝕋, *R*), where 𝒮 is a set of states, 𝒜 is a set of actions (i.e., treatments), 𝕋 is a (hidden) transition function such that 𝕋 (*s*′|*s, a*) gives the probability of transitioning from state *s* ∈ 𝒮 to state *s*′ ∈ 𝒮 given action a ∈ 𝒜, and reward function *R* is such that *R*(*s, a, s*′) is the reward received when action *a* is performed in state *s* and leads to a transition into state *s*′. In the rehabilitation case, the states are motor “memories” on which the outcomes will depend, the actions refer to the dose and type of rehabilitation at a given timestep, the transition function is equivalent to the patient (dynamics) model that describes the person’s response to treatment, and the reward function generates the reward (i.e., a positive reinforcement signal) given to the agent that quantifies the goodness of the chosen treatment at each timestep.

As a branch of AI, RL aims to design an *agent* (i.e., an autonomous decision-maker) that can learn to act optimally in an unknown “environment” commonly modeled by an MDP. The optimal actions maximize the instantaneous and total future rewards (in expectation). For precision rehabilitation, the goal of an RL agent is to learn an individualized, reward-maximizing treatment *policy* (i.e., a protocol for selecting treatments contingent on the patient’s clinical state), where the reward function is customized by the clinician and the patient based on the desired long-term outcomes. The learning process in an MDP is interactive: the agent tries different treatments, the patient generates a reward signal, and the agent adjusts its strategy intelligently to make sure that better treatments are more likely to be selected.

To address the specific challenges of precision rehabilitation post-stroke outlined in Background, we propose a novel AI-agent based on contextual model-based RL with four key elements. The RL agent1) utilizes an interpretable, dynamical, Bayesian patient model that takes into account the time-varying (dynamical) processes of stroke recovery at different time scales modulated by treatment decisions (item ➀ in Figure 1), 2) performs patient model update via contextual and hierarchical modeling (item ➁ in Figure 1), 3) takes into account constraints and personal preferences (item ➂ in Figure 1), and 4) plans a sequential treatment with respect to the context, constraints, and uncertainties in the patient model (item ➃ in Figure 1). Each component is described in the next four sections.

**Figure 1.**
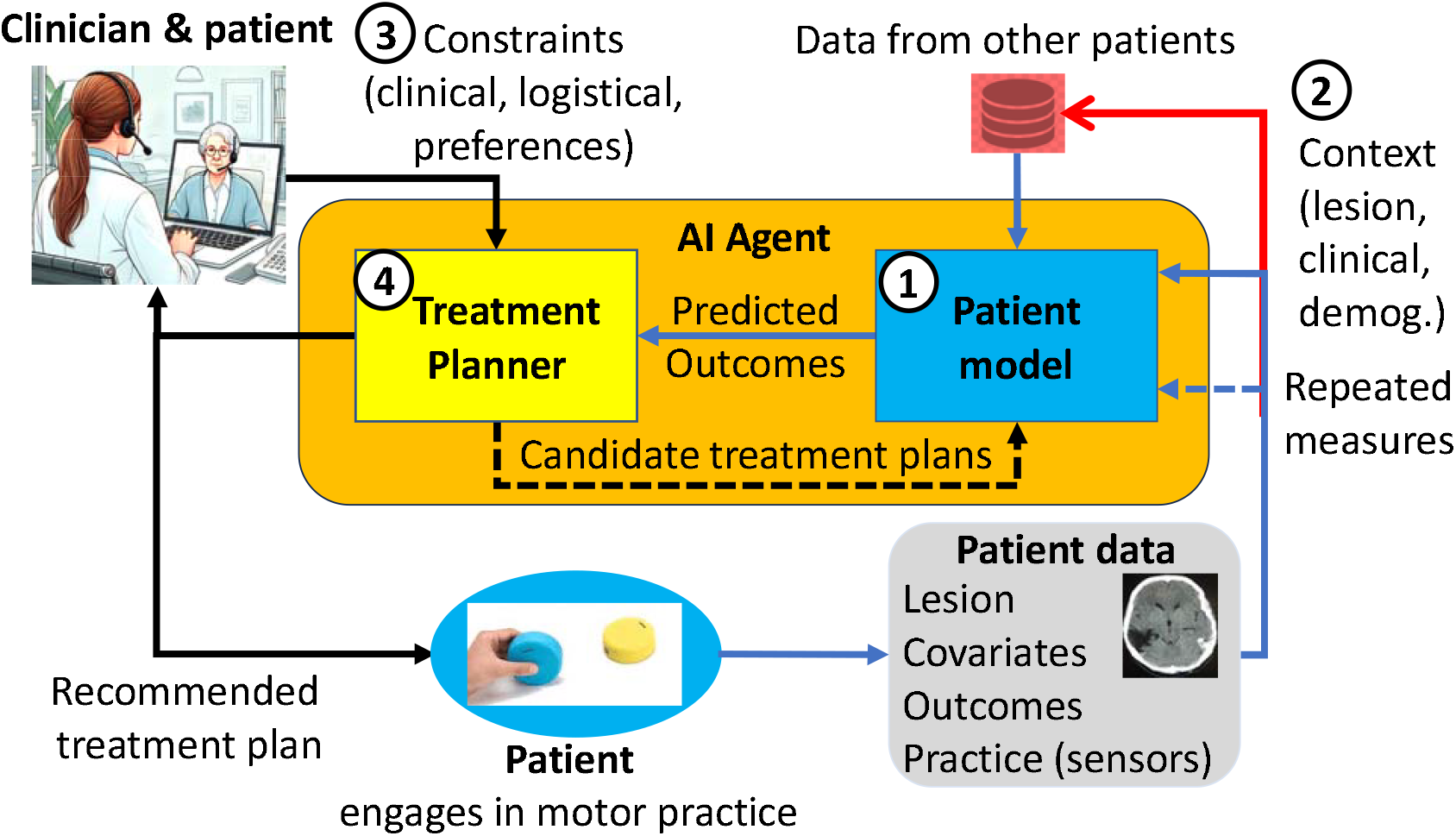
The collaborative AI precision neurorehabilitation system has four main components: (1) a patient model informed by (2) contextual data from the current and other patients, (3) constraints and preferences as determined by the clinician and the patient, and (4) a treatment planner based on PSCRL, i.e., Posterior Sampling for Contextual Reinforcement Learning. The patient model and the treatment planner form the AI agent that is capable of autonomously proposing treatment plans that maximize the predicted outcomes. Note that the image showing the “Clinician & Patient” is not an image of real people but is an artificially generated cartoon.

#### Interpretable, dynamical, Bayesian patient model for precision rehabilitation

Feasible implementation of RL for precision rehabilitation requires a patient model that forecasts (“predict some future condition as a result of study and analysis of available pertinent data”(62)) functional outcomes accurately and precisely during the subacute to chronic phases, given the current state and action. Unlike in *model-free* RL, in which a control policy is directly learned via slow trial and error, in *model-based* RL, the system learns a model of the environment (i.e., transition probabilities and reward functions) via (efficient) supervised learning and then solves the MDP using this learned model. Because model-based RL requires fewer interactions with patients to identify good policies, model-based RL is preferred in medical applications (7-10, 63). (For instance, there could be as little as one interaction between the AI system and the patient every two weeks, whereas 10,000s of interactions would be needed in a model-free approach).

A second significant advantage of model-based RL is the possibility of interpretability of the decision-making process. Although “black box” models, such as recurrent neural networks, could be used for patient modeling, mechanistic, interpretable, “grey box” models (i.e., with a structure that is motivated by theory and the parameters estimated from patient data) can be developed to explicitly account for the time-varying processes of stroke recovery, where the dosage, intensity, and timing of practice modulate recovery (see Background section). For instance, in previous work (29), we used state-space modeling to forecast UE functional outcomes for chronic stroke that included a) the intensity of practice (e.g., the daily dose, constrained by the total dose) as input to account for gain in function due to motor training, b) a “forgetting” term to account for the need of distributed practice, and c) a non-linear “self-training” term to account for the dose of UE use in daily activities. Analysis of the parameters in such interpretable models can help the clinician make informed decisions about therapy. For instance, in our previous model, if the estimated forgetting rate is high, more frequent “booster” sessions need to be scheduled. An updated model for all phases of stroke recovery, from acute to chronic, would include a critical window that modulates training effectiveness as a function of time since stroke and a spontaneous recovery term.

Finally, because the RL agent must select treatment plans that account for the uncertainty of recovery post-stroke, the patient model needs to quantify uncertainty in long-term predictions, e.g., by providing credible intervals for future outcome assessments given a treatment plan. Bayesian modeling provides a principled framework for uncertainty quantification and for incorporating prior knowledge (when available, e.g., from similar patients in a database) to reduce such uncertainties in the predictions.

#### Contextual and hierarchical modeling to leverage data from other patients

Because post-stroke response to treatment largely depends on individual characteristics (see Background), the RL agent must find individualized treatment policies. In a typical model-based RL application, such as robot control, the model is typically well-identified and identical across all instances of the controlled system (e.g., the robots). In contrast, humans show large inter-individual variations, which are further magnified by the variability of the stroke. Therefore, we propose a Contextual MDP(64) (CMDP) framework for precision rehabilitation. CMDP can be seen as a collection of (individual level) MDPs connected by contexts (lesion, etc.; Item ➁-Context in Figure 1). The context is an arbitrary set of measured covariates (e.g., type of stroke, see above) that partially explains individual differences in outcomes. CMDP assumes context-dependent dynamics (i.e., the patient’s outcomes trajectory partially depends on the context) and is thus suitable for modeling multi-patient, heterogenous rehabilitation data(65). For instance, the context can be included in the hierarchical patient model by linearly (or non-linearly) influencing the gains due to motor training.

A difficulty in forecasting neurorehabilitation outcomes, however, is that for a new patient, there is initially no (or little) data to estimate the effect of motor therapy, and the variance of the predictions may be large. As in our previous work, we therefore consider a hierarchical Bayesian model to refine the initial predictions via population level “hyper-parameters” that allows information sharing from past, similar patients (29). Crucially, the hyper-parameters are used to construct informed prior distributions for individual level parameters when predicting the response of a patient early in therapy. This hierarchical model is trained with repeated data from sensors and clinical assessments, baseline contextual data from the current patient and an expanding patient database (see Figure 1).

#### Treatment constraints and preferences limit the range of possible treatments

Treatment plans must take into account clinical and logistical constraints as well as personal scheduling preferences (Item ➂ in Figure 1). Thus, we consider finding an optimal constrained treatment policy in a CMDP with respect to, for instance, realistic constraints such as a long-term rehabilitation budget (i.e., the total dose that can be administered throughout the longitudinal treatment), a stepwise dose limit (e.g., at most two therapy sessions per week). This leads to a novel constrained RL problem in a CMDP with unknown individualized dynamics. Most constraints and preferences (e.g., a dose limit per week and scheduling restrictions due to time conflict) can be handled through a time-varying action set, which reduces the search space for an optimal treatment plan. However, special care is required for an RL algorithm to strictly adhere to the long-term constraints (e.g., the total dose). In this work, we propose a budgeted dynamic programming method that plans according to both the patient’s state and the remaining budget to solve the long-term planning problem.

Via the collaborative nature of our AI system (see Figure 1), the clinician and patient can input “hard” constraints, such as the minimum or maximum daily distal and proximal arm practice doses every two weeks, and, if needed, “soft” constraints, such as the weights of the reward function. For instance, in preliminary simulations (see below), the reward is the sum of the outcomes at 6 months plus the mean outcome until then with equal weighting. This can be adjusted, for instance, for more emphasis on long-term outcomes or specific treatment goals (i.e., emphasis on distal vs proximal arm functions). After the adjustments, the algorithm will be rerun, and the clinician will be able to visualize the proposed plan and forecasted outcomes and modify it as desired.

#### A treatment planner that accounts for uncertainty in the patient model, context, and constraints

We consider the RL problem in CMDP, focusing on the uncertainties of the individualized model and the update of this model. We propose a novel self-improving treatment planner (Item ➃, in Figure 1) based on the Posterior Sampling for Contextual Reinforcement Learning (PSCRL) algorithm for solving the RL problem in CMDP. PSCRL is a model-based algorithm that tackles the challenges of precision rehabilitation by combining Bayesian hierarchical modeling and planning under constraints. PSCRL belongs to a family of algorithms following Thompson sampling (66, 67) (68), which exhibits strong empirical performance and theoretical guarantees in various RL settings, including recent medical applications (69-72). Briefly, at each step, PSCRL updates a posterior distribution over individual level MDPs, takes one sample from this posterior, and optimizes treatment for this specific patient model. This posterior-sampling behavior tackles the tradeoff between exploration and exploitation. Early in learning, as the model is uncertain, with wide parameter distributions, rehabilitation plans are varied. Late in learning, as uncertainty decreases, the plans are closer to optimal. Once a specific model instance is selected via sampling, we apply a planning algorithm (such as dynamic programming in our simulation study below). To account for budget constraints, our treatment plans select treatments with respect to both the current patient state and the remaining budget. In the following, we describe PSCRL in the context of stroke rehabilitation.

### Formal description of the RL algorithm for dose scheduling

To illustrate the functioning of our AI-based precision rehabilitation system, we introduce the CMDP formulation for a simple dose optimization problem of a generic upper extremity treatment with finite dosing options, which represents the hours of rehabilitation therapy with realistic scheduling constraints. We defer the technical details to Supplementary Material A. We then describe the PSCRL algorithm to solve this problem.

#### Notations

We first define necessary notations for formalizing the sequential dose optimization problem. For any positive integer *m*, we define [*m*] = {1,2,…}. For any two positive integers *n* ≤ *m*, we use the the shorthand *n* : *m* = {*n, n* + 1, …, *m*}. We denote an ordered collection of values (or random variables) by applying this notation in subscript. For instance, we write *s*_*i*,1:*H*_ = (*s*_*i*_,_1_,*s*_*i*_,_2_…,*s*_*i*_,_*H*_) to denote a sequence of states for patient *i* ∈ ℕ until timestep *H* ∈ ℕ. For *d* arbitrary real numbers *a*_1_,*a*_2_,…,*a*_*d*_, we also use *a*_1;*d*_ = (*a*_1_,*a*_2_,…, *a*_*d*_) to denote a *d*-dimensional vector, i.e., *a*_1;*d*_ ∈ ℝ^*R*^.

### Description of the Precision Rehabilitation scenario

We consider a scenario in which a clinician-AI team treats, in sequence, a cohort of *N* patients post-stroke. Each patient receives rehabilitation treatments over a fixed treatment horizon (e.g., 6 months) containing *H* discrete timesteps (e.g., each timestep is two weeks). Upon patient intake, the clinician-AI team observes a *d*_*c*_-dimensional context 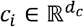 that encodes *d*_*c*_ clinical covariates (demographic information, type of stroke, etc.) for the patient indexed by *i*. Then, at time *t* ∈ [*H*], a patient’s recovery is measured via clinical outcome assessments summarized into an outcome *o*_*i,t*_ ∈ 𝒪 ⊆ ℝ (e.g., the Action Research Arm test, ARAT, or the Motor Activity Log, MAL), where 𝒪 is called an outcome space; for instance, 𝒪 = [0,5] for the MAL. We assume that the outcome *o*_*i,t*_ depends on a latent state *s*_*i,t*_ ∈ 𝒮 ⊆ ℝ that summarizes the patient’s motor state (or “memory”) at time *t*. We consider scalar outcome and treatment for ease of demonstration. We discuss vector-valued outcomes and treatments (e.g., a 2-dimensional dose that specifies distal and proximal practice separately) in **Discussion: Future Work**.

For each patient *i* and each timestep *t*, the collaborative AI system recommends a treatment *a*_*i,t*_, which represent, for instance, the hours of rehabilitation therapy in this simple dose optimization example. The treatment influences the motor state and in turn the outcome in future timesteps. To reflect realistic time and monetary constraints, we also impose a total budget of 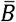 therapy hours to be distributed over *H* timesteps (i.e.,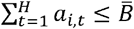) and a step-wise limit 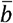 that represents the maximum rehabilitation dose that can be administered at a timestep (i.e.,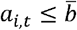). These constraints can be modified by the clinicians or the patients at any time.

### Data generating process of multi-patient rehabilitation data

We compactly denote the data collected after treating patient *i* by a context-trajectory pair (*c*_*i*_, τ_*i*_) where τ_*i*_ = (*o*_*i*,1:*H*+1_,*a*_*i*,1:*H*_) is a trajectory containing the history of outcomes and treatments throughout the *H*-step treatment horizon, with an additional post-treatment outcome *o*_*i,H*+1_. In general, we assume that the outcome *o*_*i,t*_ only partially reveals the patient’s latent state *s*_*i,t*_(e.g., motor memory) and is a random variable that is conditionally independent of all other variables given *s*_i,t_. We represent this dependency by an observation function 𝕆(*o*∣*s*), which gives the probability of observing outcome *o* when the current patient-state is *s*. Here, the observation function 𝕆 is stochastic to reflect random measurement errors of clinical assessments. We write *o*_*i,t* ∼_ 𝕆(· ∣ *s*_*i,t*_)to denote that the conditional distribution of *o*_*i,t*_ is 𝕆(· ∣ *s*_*i,t*_). We further assume that for each patient *i*, states *s*_*i*,1:*H*+1_ and treatments *a*_*i,1:H*_ follow a first-order dynamics model parametrized by an unknown patient-specific *d*_*θ*_-dimensional vector 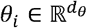 Specifically, for each *t* ∈ [*H*], given the current state *s*_*i,t*_ and treatment *a*_*i,t*_, the next state *s*_i,t+1_ follows a conditional distribution 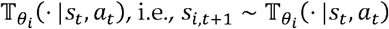.

In addition, the context vector *c*_*i*_ observed at intake can be used to infer the unknown parameter vector θ_*i*_ that characterizes the transition dynamics. Components of *c*_*i*_ are covariates that potentially encode similarity between patients (see Background section) and can be used to inform clinical decision-making when little to no outcome data is available for a new patient *i*. We assume that the exact relationship between contexts and recovery dynamics is unknown to the clinician-AI team and needs to be inferred from data. A reward signal is defined based on observed data.

In summary, the interactions between the clinician-AI team and the patients can then be described with the following:

1. For patient *i* = 1,2,…, *n:*
2. Conduct baseline measurements on patient *i* to observe context 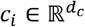
3. At timestep *t* = 1,2,…,*H*:
4. Observe patient outcome *o*_*i,t*_ ∼ 𝕆(· ∣ *s*_*i,t*_) and remaining budget *b*_*i,t*_
5. Clinician-AI team decides and recommends dose 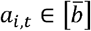 subject to constraint *a*_*i,t*_ ≤ *b*_*i,t*_ and user-defined rewards, using data from past patients (*c*_1:*i*-1_, τ_1:*i*-1_) and from the current patient (*c*_*i*_,*o*_*i*,1:*t*,_*a*_*i*,1:*t*-1_)
6. With treatment *a*_*i,t*_, patient undergoes transition in (latent) state, 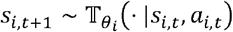 budget updates to *b*_*i,t*+1_ = *b*_*i,t*_ – *a*_*i,t*_
7. At timestep, the patient returns to the clinic for post-treatment measurements, leading to terminal outcome *o*_*i,H*+1_

Unlike an MDP, this scenario assumes that the patient-state *s*_*i,t*_(e.g., motor memory) is hidden from the clinician-AI team and the outcomes are generated by an observation function 𝕆. In theory, this environment is called a Partially Observable MDP (POMDP), which is computationally intractable in general. In practice, RL in POMDP can be approximately solved by expanding the state with measurements from multiple timesteps (see Discussion). For simplicity, in the rest of the paper, we consider the MDP case with fully observed states and the trajectory for a patient becomes *τ*_*i*_ = (*s*_*i*,1_,*a*_*i*,1_,…,*s*_*i,H*_,*a*_*i,H*_,*s*_*i,H*+1_).

#### Algorithm: Posterior Sampling for Contextual Reinforcement Learning (PSCRL)

Here, we briefly describe the PSCRL algorithm (Algorithm 1).^b^ At time *t* for patient *i*, PSCRL first updates the posterior distribution of parameters 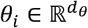 of the patient model given all available data. As a shorthand, we denote the posterior density of θ_*i*_ by

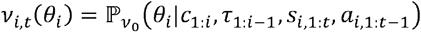

where *ν*_0_ is a prior distribution over all unknown variables and 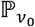 indicates that the posterior distribution depends on the prior *ν*_0_. Then, a plausible parameter vector 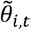 is randomly drawn from this posterior distribution over the model parameters for the current patient. Next, an integer-valued dose 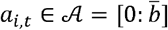 subject to user-specified constraints is determined by an optimal control algorithm (such as dynamic programming used in the simulation study below) on the sampled dynamics model. Specifically, PSCRL computes a time-dependent optimal treatment policy 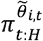 for the remaining timesteps such that 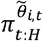 maximizes the expected predicted future rewards, i.e.,

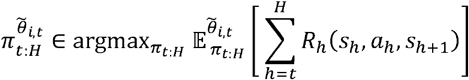

where 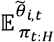 indicates that the expectation is taken over the distribution of future states and actions under policy *π*_*t*:*H*_ (i.e., *a*_*h*_ = *π*_*h*_ (*s*_*h*_) for *h* = *<u>t,t</u>*+ 1,…, *H*) and the sampled dynamics parameter 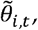 and *R*_*t:H*_ is the user-defined, time-varying reward function (which may be modified at will, e.g., to reflect the desired long-term rehabilitation outcomes by the clinician-patient team). According to this (updated) policy, the recommended treatment at timestep by PSCRL is. As new outcome data is observed in the next timestep, the posterior distribution is updated, leading to more accurate (sampled) patient models, better treatments, and better outcomes.

##### Algorithm 1 Posterior Samoling for Contextual RL (PSCRL)

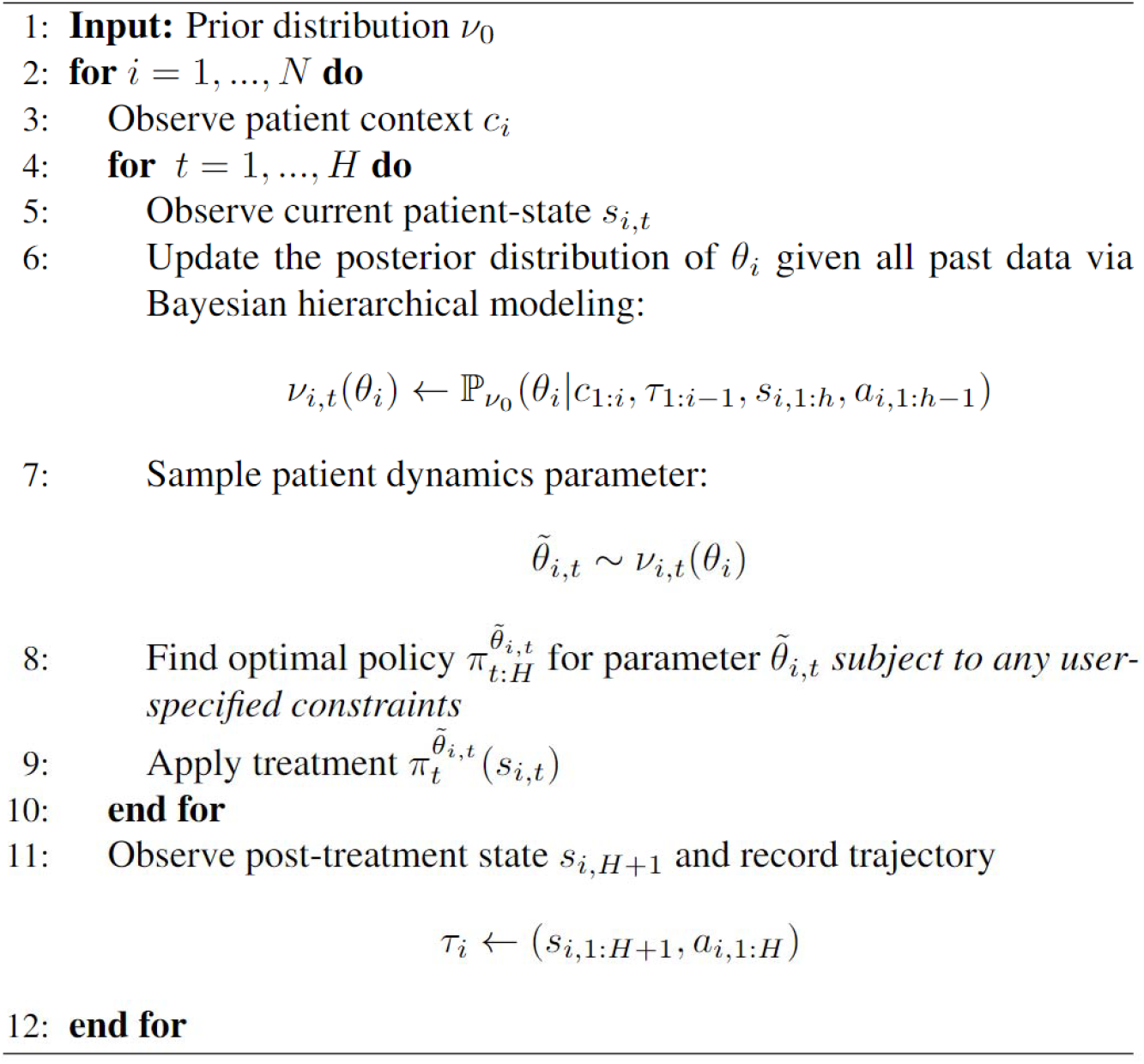

### Simulation study: adaptive dose scheduling in chronic stroke

We tested our new PSCRL algorithm in simulations using a chronic stroke model. In previous work(29), we developed and validated a chronic model based on DOSE and EXCITE clinical trials (20, 73), which provided compelling evidence that the dose and scheduling of therapy affect rehabilitation outcomes for patients with chronic stroke. As a testbed for the proposed framework, we developed a simulator based on our previous dynamics model for the change in the Motor Activity Log (MAL) (29), a functional UE measure with slight modifications to include a patient context (see below). We simulated *N* = 100 patients arriving in sequence to receive rehabilitation therapy over a 6-month treatment window. The treatment horizon was divided into *H* = 12 timesteps, where each timestep corresponds to a 2-week interval. At each timestep, a patient may receive a dose of 0—20 hours of rehabilitation therapy, with a maximum total dose of 60 hours, corresponding to the maximal dose and the maximal weekly dose, respectively, in the DOSE trial(20). Large between-patient variability was modeled by including four baseline covariates.

#### Specifications of the patient simulator

##### Patient dynamics model

Henceforth we use superscript “⋆” to denote a ground-truth parameter used by the simulator but hidden from the decision-maker. Following our previous work(29), our simulation utilizes a patient dynamics model (or patient model, for short) that describes the change in the MAL *o*_*i,t*_ ∈ [0,5] (see Table 1). Formally, the dynamics model contains a subject-independent (stochastic) observation function 𝕆(*oi,t* | *χ*_*i,t*_) that gives the conditional probability of *oi*_,*t*_ given “motor memory” *x*_*i,t*_ ∈ ℝ. The motor memory is updated by an individualized transition function 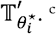. ^c^Up to some process noise, the next motor memory *x*_*i,t*+1_ is a function of the current memory 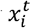 modulated by the retention rate 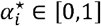, the dose of training *a*_*i,t*_ modulated by the learning rate 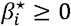, and the current MAL *o*_*i,t*_ modulated by the self-training rate 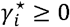. We represent individual level parameters using a vector 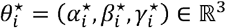. Additionally, the individualized dynamics depend on population level parameters, including a process noise scale 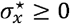, slope-and-offset parameters (*κ*_slope_, *κ*_offset_) for the observation function that converts motor memory into an MAL measurement (potentially with an observation error *∈*_*i,t*_), and a random effect scale 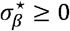 on learning rate. We update the previous model to reflect the large between-patient variability seen in stroke recovery, by assuming that the patient’s context vector *ci ∈ ℝ*^*d*^ influences 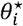 linearly via a weight matrix 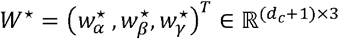, where 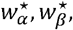 and 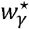 contain fixed intercepts (at first coordinate) and fixed effects. For simplicity, we only added random effect for learning rates 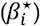.

**Table 1.**
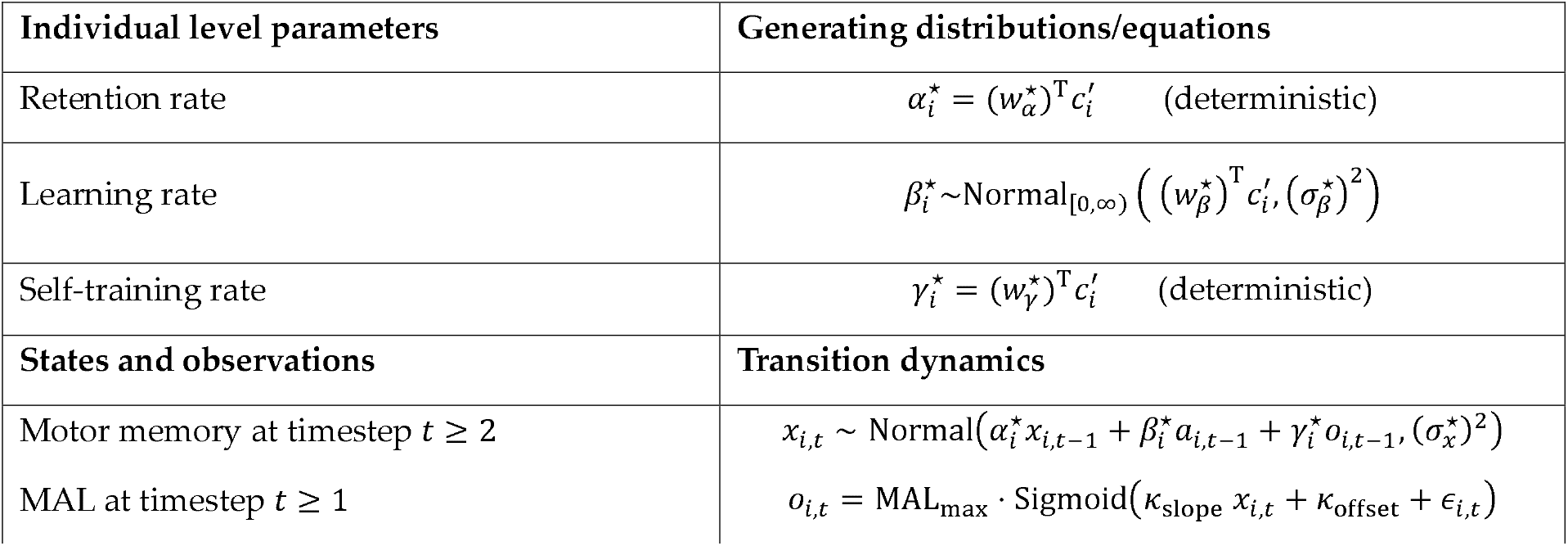
Simulation of the change in the Motor-Activity-Log (MAL) for synthetic patient in the simulation. The MAL ranges from 0 to MAL_max_ = 5. We let 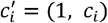 where *c*_*i*_ is the context vector for patient *i*. We assume no observation noise, i.e., ∈_*i,t*_ = 0. The initial MAL *o*_*i*,1_ ∈ [0,5]. was generated according to Normal_[0.5]_, (*μ* = 2, *σ*^2^ = 0.04), i.e., a truncated normal distribution on the interval [0, 5] The initial motor memory *x*_*i*,1_ ∈ ℝ is obtained by the inverse of the (deterministic) observation function.

| Individual level parameters | Generating distributions/equations |
| --- | --- |
| Retention rate | $\alpha_i^* = (w_\alpha^*)^T c_i'$ (deterministic) |
| Learning rate | $\beta_i^* \sim \text{Normal}_{[0,\infty)} \left( (w_\beta^*)^T c_i', (\sigma_\beta^*)^2 \right)$ |
| Self-training rate | $\gamma_i^* = (w_\gamma^*)^T c_i'$ (deterministic) |
| States and observations | Transition dynamics |
| Motor memory at timestep $t \geq 2$ | $x_{i,t} \sim \text{Normal}(\alpha_i^* x_{i,t-1} + \beta_i^* a_{i,t-1} + \gamma_i^* o_{i,t-1}, (\sigma_x^*)^2)$ |
| MAL at timestep $t \geq 1$ | $o_{i,t} = \text{MAL}_{\max} \cdot \text{Sigmoid}(\kappa_{\text{slope}} x_{i,t} + \kappa_{\text{offset}} + \epsilon_{i,t})$ |

##### Reward definition

To find a balance between the patient’s UE function during the treatment as well as the overall rehabilitation function measured post-treatment, we defined the return (i.e., total reward) for treating a patient to be the sum of the terminal MAL at 6 months (*o*_*i,H*+1_) and the mean MAL 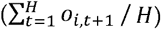 after the first treatment session. Hence, we defined the (time-varying) reward function at each step by:

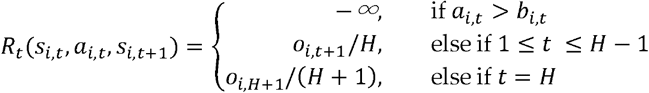

where *s*_*i,t*_ = (*o*_*i,t*_,*b*_*i,t*_) is an expanded state for the CMDP, and we use a penalty of − ∞ to encode the constraint that each dose *a*_*i,t*_ may not exceed the remaining budget *b*_*i,t*_ at the current timestep.

##### Simulation hyper-parameters

We consider *N* = 100 patients who arrive in sequence to receive rehabilitation treatment over *H* = 12 timesteps. A synthetic patient is indexed by *i* ∈ [*N*] and represented by a four-dimensional context vector *c*_*i*_ ∈ ℝ ^4^ along with an unknown (ground-truth) parameter vector 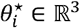. The patient contexts were drawn independently from a population distribution (specified in Supplementary Material B) with two continuous covariates (that could represent baseline function, sensory integrity, etc.) and two categorical covariates (that could represent stroke type, side affected, etc.). Dynamics parameters {*θ*_*i*}*i*∈[*N*}_ were then randomly generated by a conditional distribution given the contexts as shown in Table 1 with an unknown random effect scale 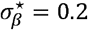 for learning rates. Other (hidden) hyper-parameters 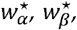 and 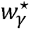 for linear context-to-dynamics relationship are specified in Supplementary Material B. For simplicity, we assumed that the observation function 𝕆 is deterministic (i.e., *∈*_*i,t*_ = 0) and known by the decision-maker with parameters κ_slope_ = 0.2 and κ_offset_ = −3. As a result, the simulator matches the assumptions of CMDP and avoids intractability issues (see POMDP in discussion). See additional details in Supplementary Material B.

#### Implementation details of the PSCRL algorithm

Implementing PSCRL requires specifying a hierarchical Bayesian model and a planning algorithm that finds the optimal policy given a sampled patient model.

##### Patient model update via posterior inference

We make the simplifying assumption that the structure of the hierarchical MAL model described above is known except for the ground-truth parameters. PSCRL uses a hierarchical Bayesian modeling approach, treating all unknown quantities as random variables. For clarity, consider the parameters with superscript “⋆” (e.g., 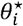) as fixed unknown quantities, while those prior without superscript (e.g. *θ*_*i*_) as random variables. The hierarchical Bayesian model is defined by hyper-prior distributions for population level random variables (or vectors), (*w*_*α*_,*w*_*β*_,*w*_*γ*_, *σ*_*β*_, *σ*_*x*_) prior distributions for individual level variables *θ*_*i*_ = (*α*_*i*_, *β*,_*i*_ *γ*_*i*_) conditioned on the population-level parameters and a likelihood function for the observed data (*o*_*i,t*_′s along with *a*_*i,t*_′s). **Table 2** shows the prior and hyper-prior distributions used by PSCRL in the simulation experiment. The likelihood function is given by the transition function as in **Table 1**. Since exact posterior inference is computationally intractable, we used Hamiltonian Monte Carlo (HMC)(74), a state-of-the-art Markov Chain Monte-Carlo (MCMC) algorithm (implemented in NumPyro (75)), to approximate the posterior distribution. MCMC algorithms are generally considered “exact” posterior inference algorithms since the approximation error can be arbitrarily small when the number of samples from the posterior distribution is large.

**Table 2.** Bayesian model for unknown parameters in the individualized dynamics. “Priors” are the prior distributions for the individual level parameters conditioned on population level parameters. “Hyper-priors” specify the prior distributions for population level parameters. The process noise scale (*σ*_*x*_ is shared across patients and is used to compute the likelihood of outcome trajectories according to the transition dynamics in Table 1. Known parameters are omitted. The (4-dimensional) zero-vector is denoted by **0** and the (4-by-4 identity matrix is denoted by ***I***.

| Unknown parameters | Priors | Hyper-priors |
| --- | --- | --- |
| Retention rate | $\alpha_i = w_\alpha^T c_i'$ (deterministic) | $w_{\alpha,0} \sim \text{Normal}(0, 1)$<br>$w_{\alpha,1:4} \sim \text{Normal}(\mathbf{0}, 0.1\mathbf{I})$ |
| Learning rate | $\beta_i \sim \text{Normal}_{[0,\infty)}(w_\beta^T c_i', \sigma_\beta^2)$ | $w_{\gamma,0} \sim \text{Normal}(0, 1)$<br>$w_{\beta,1:4} \sim \text{Normal}(\mathbf{0}, 0.1\mathbf{I})$<br>$\sigma_\beta \sim \text{HalfNormal}(0.5)$ |
| Self-training rate | $\gamma_i = w_\gamma^T c_i'$ (deterministic) | $w_{\gamma,0} \sim \text{Normal}(0, 1)$<br>$w_{\gamma,1:4} \sim \text{Normal}(\mathbf{0}, 0.1\mathbf{I})$ |
| Process noise scale | | $\sigma_x \sim \text{HalfNormal}(1)$ |

##### Planning for optimal treatment policy under constraints

Given a sampled parameter 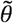, PSCRL solves the associated constrained planning problem subject to a total rehabilitation budget 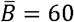 and a. stepwise dose limit. The dose limit 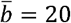. The dose limit restricts the MDP’s action space to 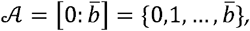 which simplifies the planning problem. Handling the budget constraint requires long-term planning, as choosing a dose now may affect dosing options in the future. A simple solution for this is to consider an MDP with an expanded state space 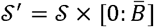 with deterministic transitions in the remaining budget component(76)]. Then, an optimal policy for the expanded MDP is equivalent to an optimal constrained policy for the original MDP. Although the state space for the MAL model is continuous, in this simulation example, we approximately solve the constrained planning problem using Dynamic Programming (DP) with the expanded state space and discretized MAL outcomes (100 bins of width 0.05 covering the range of MAL [0.,5]).

#### Evaluation protocol

##### Regret (performance metric)

To rigorously examine the potential benefit of AI-based precision rehabilitation, we measured the performance of a learning agent by a “regret” metric (similar to (64)). In this work, we define the *regret* of an RL agent after seeing *N* patients as:

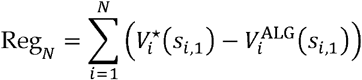

where *s*_*i*,1_ is a fixed (pre-generated) initial state for patient *i*, 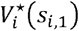 is the expected return of a *theoretically optimal policy* associated with patient *i* starting at *s*_*i*,1_, and 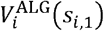 is the expected return for treating the same patient using treatments recommended by the algorithm. The expectation (in expected return) is taken over the randomness of state transitions (i.e., process noise) and treatments. A lower regret is preferred as it suggests that (in expectation) the overall treatment effect is closer to the theoretically optimal effect. Because patients are heterogenous (represented by different parameters 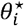), the optimal treatment policy and its corresponding value vary across patients. Due to infinite state space, non-linear(stochastic) transitions, and constraints, both 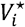 and 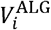 are impossible to compute exactly. We thus estimated 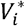 with a near-optimal benchmark obtained using dynamic programming with access to the ground-truth parameter 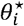. Notice that 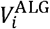 also depends on the randomness of the algorithm (which in turn depends on all data from past *i* − 1 patients). We estimated 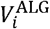 by averaging over the results from 10 independent runs of the algorithm.

##### Comparison with non-adaptive treatment (benchmarks)

We compare the regret performance of PSCRL against three non-adaptive benchmark treatment policies: Uniform, Increasing, and Decreasing. For any patient, Uniform distributes doses evenly throughout the treatment horizon while Increasing and Decreasing administer doses in an increasing and decreasing manner in fixed steps (respectively).

##### Comparison of PSCRL with and without within-patient learning

By updating the patient model using both covariate data at baseline and all measurement data during training, PSCRL is self-improving both at the patient and population levels. That is, as the database of past patients expands, PSCRL can recommend treatment policies that are increasingly effective in improving functional gains for any new patient, while continuously improving the policy by finetuning the (Bayesian) model with incoming patient-specific data over time and across the patient population. To test the effect of individual level learning when new outcome data becomes available, we investigated the performance of a simplified version of PSCRL (dubbed PSCRL-reduced) that only constructs a patient model and plans accordingly once at the intake stage for every patient. In other words, there is no within-patient learning and replanning in the reduced-PSCRL.

##### Fixed patient sequence

Notice that in the CMDP formulation, the expected return of an agent (or a treatment policy) may vary significantly between patients due to different individualized transition functions and different initial states. Hence, to avoid unnecessary variations in the performance metric, we evaluated the agent and the benchmark treatments with a fixed sequence of synthetic patients. The patient sequence is generated in advance by the simulator according to the data-generating process specified in Supplementary Material B.

##### Average return on the population level

We further measured the performance of an agent (or a treatment policy) by its average return when treating the patient population. The *average return* was evaluated by averaging over the expected return of the agent (or a policy) on 50 held-out patients. The held-out patients were pre-generated from the same distribution as the patients occurred during the learning stage. This metric is particularly useful to evaluate the performance of an RL agent with different levels of simulated clinical experience (i.e., number of past patients in its database).

## Results

### Evidence of effective, personalized treatments with step-by-step replanning

In Figure 2, we illustrate the model predictions and treatment policies proposed by PSCRL for two synthetic patients along with the history of the posterior distribution of the learning rate *β*_*i*_. progression of the predicted trajectories is presented in Fig2A-C (light blue) for timesteps *t* = 1,7,13 (respectively). Because at the initial timestep (*t* = 1, shown in Figure 2A), the individual level posterior was based on the data from past patients only, the posterior on *β*_*i*_ (shown in Figure 2D) was wide and the estimated distribution of future predicted outcomes (following the current policy 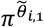) deviate rather significantly from the ground-truth (light orange in Fig 4A-B), indicating a lack of knowledge on the current patient. By learning from incoming patient data, PSCRL quickly improves its estimate of the patient model, as reflected by a narrower posterior on and more accurate predictions (as shown in Figure 2B) by the midpoint of the treatment horizon (). As a result of an improved patient model, PSCRL’s treatment recommendations became increasingly effective, leading to higher (expected) returns (see patient indices in top right of each panel) for treating both patients.

**Figure 2.**
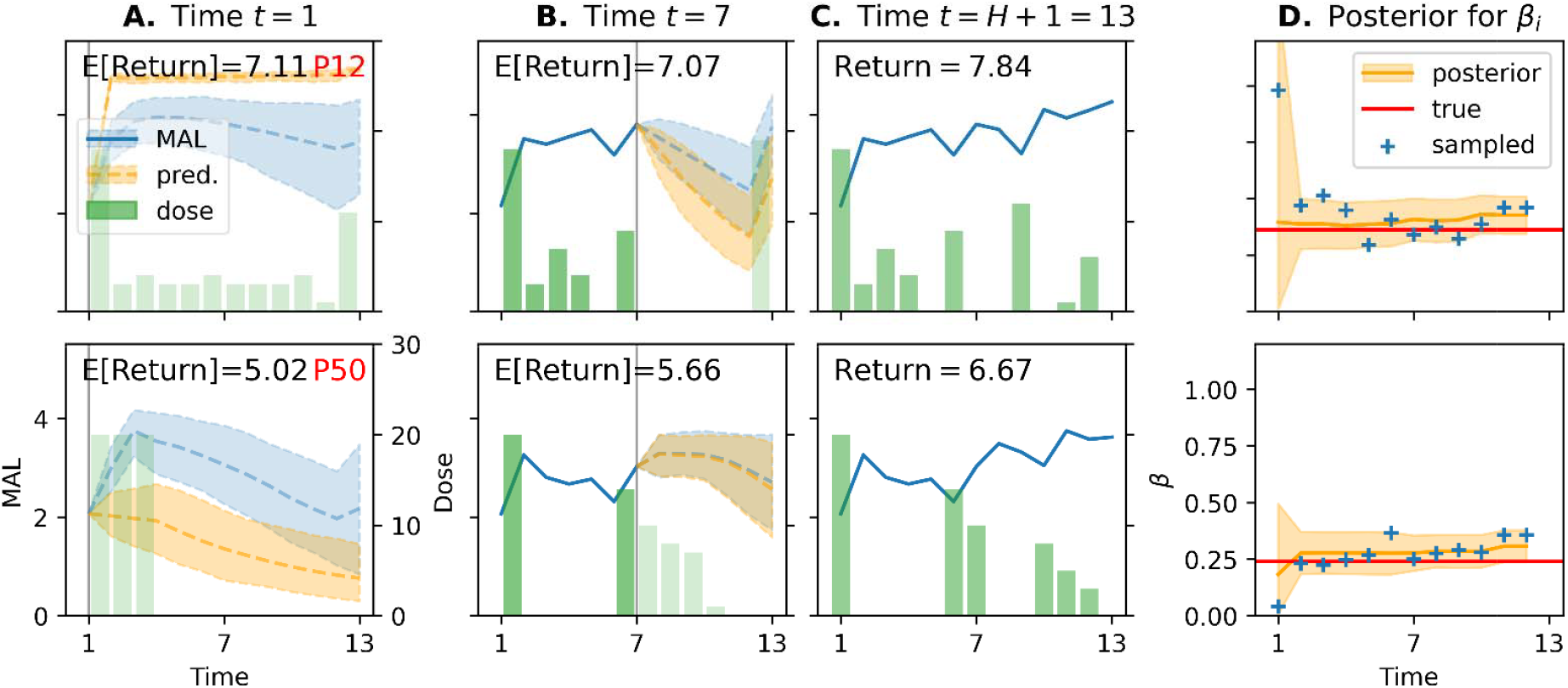
Simulation of two patients illustrating updates of model predictions and recommended treatment plans from PSCRL. **A-C. Observed, predicted, and potential (future) trajectories at initial (A), midpoint (B), and post-treatment (C) timesteps. Solid blue line**: Observed MAL outcomes. **Light blue**: Potential future outcomes with means (dashed line) and 95% confidence intervals (shaded) as generated by the true model with parameter assuming that the patient would follow policy. **Light orange:** Predicted future outcomes with means (dashed line) and 95% prediction intervals (shaded) as generated by the patient model and policy. **Solid green bars:** Past (actual) treatments. **Light green bars:** Example of future treatments assuming the patient follows (without updates from PSCRL). Notice how future treatments change as PSCRL continuously refines its policy with the new patient data. **Vertical gray line:** Indicator of the current timestep. **D. Distribution of the learning rate parameter (see Table 2) as a function of time during treatment. Orange:** Posterior distribution (mean and 95% credible interval) of the learning rate, as estimated by PSCRL at each timestep. **Blue cross:** Sampled parameter used for planning by PSCRL. **Red:** True parameter.

### Improved average return with a larger database

Figure 3 shows the performance of PSCRL with and without within-patient learning with respect to the number of past patients in the database. The database consisted of contexts and trajectories (*c*_1:*n*_, *τ*_1:*n*_)from past *n* patients. PSCRL’s adaptive treatments consistently outperformed the Uniform treatment policy even when the database size is small. Note that, in these simulations, the improvement in performance over time was small as the performance was already close to the theoretically optimal value at the beginning of the experiment thanks to individual level learning and replanning (i.e., policy updates) at each time step. In contrast, the reduced PSCRL with no within-patient learning and replanning initially showed worse performance than the non-adaptive *Uniform policy*. However, after interacting with about 10 synthetic patients, the performance of reduced PSCRL clearly improved and became consistently better than the *Uniform policy*. This result clearly shows the algorithm’s ability to learn and generalize the information between patients in the proposed setup.

**Figure 3.**
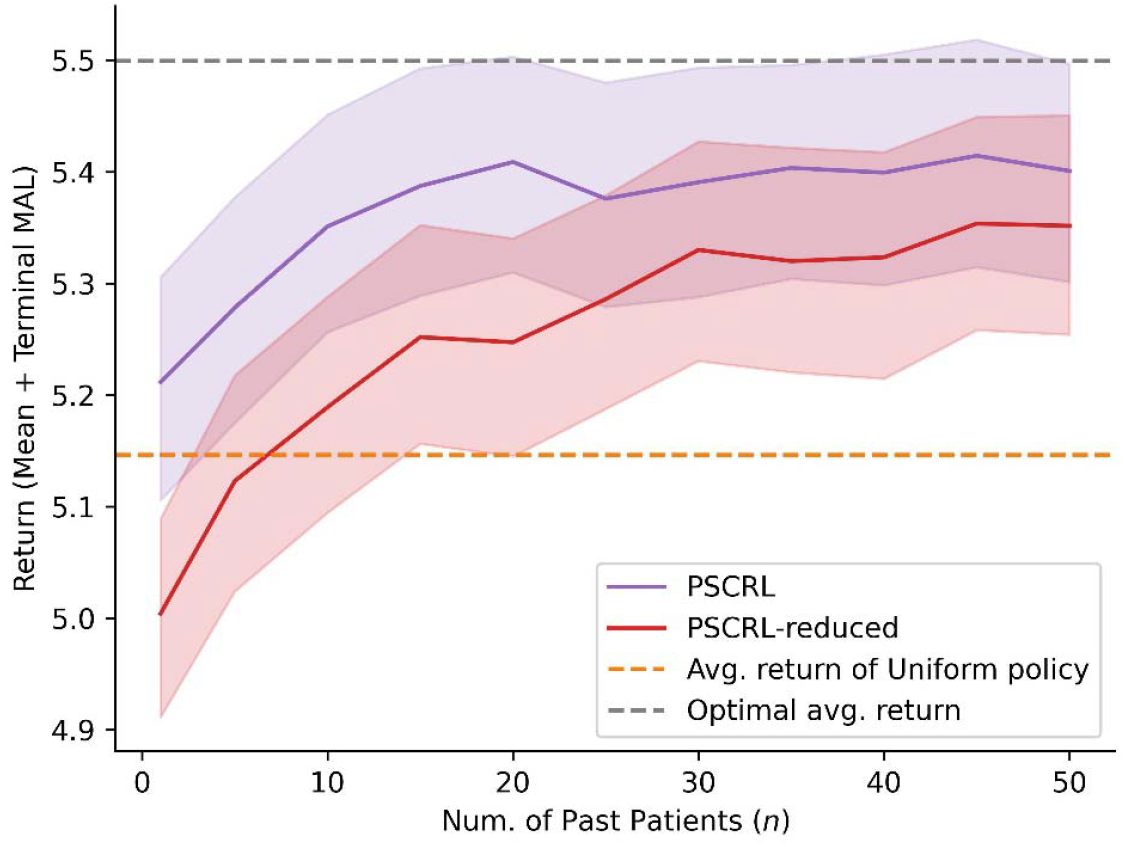
Average return of PSCRL with increasingly larger databases compared to benchmarks. **Purple:** Average return (solid) with 95% CI (shaded) for PSCRL. **Red:** Average return (solid) with 95% CI (shaded) for PSCRL-reduced (which does not perform within-patient learning). Average returns for PSCRL and PSCRL-reduced were computed by first computing the expected return of the agent (with different numbers of past patients in the database) with 40 simulations on each held-out patient and then by taking the average over all 50 held-out patients. **Orange (dashed):** Average return for *Uniform* benchmark treatment policy. **Grey (dashed):** Optimal average return, computed by averaging over the expected return of individualized optimal policies (obtained using ground-truth parameters). Expected returns for *Uniform* and optimal policies were estimated using 1000 simulations.

We then tested how well PSCRL could estimate the context parameters linking each covariate to the main model parameters in the state space model. As discussed above, the exact relationship between contexts and recovery dynamics was unknown to the clinician-AI team and had to be learned from sequential patient data (as each patient is associated with a single measurement for each covariate). As shown in Supplementary Material C, Figure S1, the posterior distribution converges to the true values, showing that PSCRL achieves learning on the population level via accurate estimation of the relationships between contextual covariates and patient parameters.

### Overall expected gain with AI-determined treatments

The overall quality of the treatment is measured by the regret (see above). Figure 4 clearly shows the superiority of PSCRL’s treatments against the benchmark treatments in minimizing regret after treating the same sequence of *N* = 100 patients. The benchmarks correspond to non-adaptive, “one-size-fits-all” dosing schedules and thus incur high regret when treating a heterogenous patient population. In contrast, even without step-by-step replanning, PSCRL-reduced learns to incur low regret, which indicates that it can still identify high-quality treatment policies for new patients by leveraging the context. The full PSCRL algorithm incurred the least regret, indicating that policies proposed by PSCRL were the closest to a hypothetical clinician with perfect information about each patient a priori.

**Figure 4.**
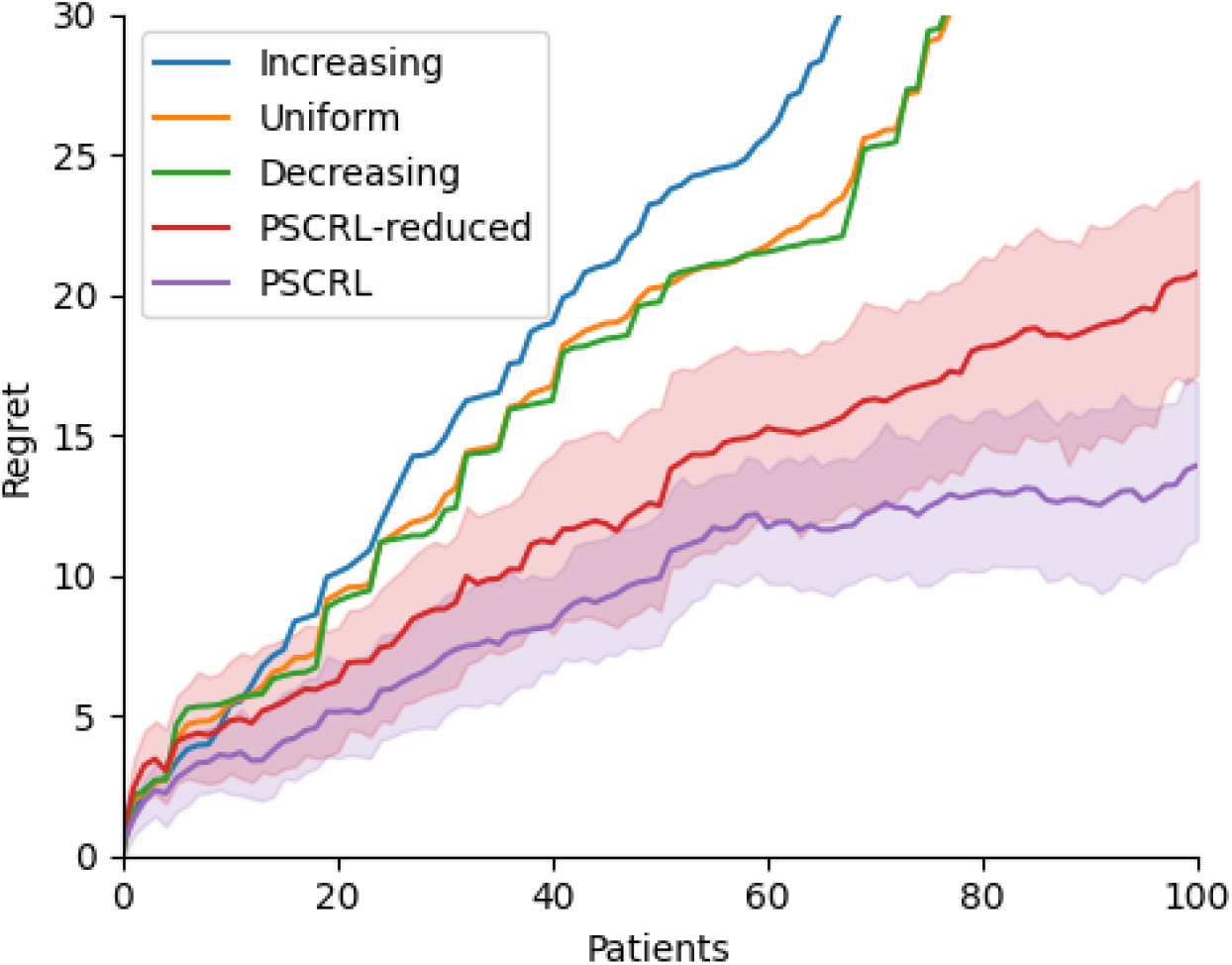
Comparison of cumulative regret for PSCRL and benchmark treatments. **Solid lines:** (cumulative) regret, estimated from results of 10 independent simulations for an RL agent or 1000 simulations for a benchmark treatment policy. **Shaded region:** 95% CI of the regret (omitted for benchmarks for readability). Given the horizon and the total budget therapy hours. *Uniform* allocates therapy hours at each timestep for any patient (regardless of the patient’s state). Decreasing sets,, and therapy hours (and 0 dose when is even) for any patient. *Increasing* uses the reversed dose schedule of *Decreasing*.

## Discussion

Determining the optimal sequential treatment plan that maximizes long-term outcomes for each patient post-stroke is daunting even for experienced clinicians because of the multiple scheduling factors that modulate recovery, the high between-patient variability, and the multiple scheduling constraints and patient preferences. Thus, to enhance the effectiveness of rehabilitation, we proposed a collaborative AI precision rehabilitation system that uses model-based RL. The system addresses challenges specific to precision rehabilitation system by 1) utilizing an interpretable, dynamical, Bayesian patient model that takes into account the time-varying processes of stroke recovery at different time scales modulated by treatment decisions, 2) updating the patient model via contextual and hierarchical modeling, 3) collaborating with clinicians and patients to customize the recommended plan based on clinical judgment, constraints, and preferences, and 4) planning a sequential treatment with respect to the context, constraints, and uncertainties in the patient model. To design such a system, we extended current medical RL applications based on MDP (7-10, 63, 77) and bandit models (equivalent to a 1-step MDP) (69, 78), and formalized precision rehabilitation as a sequential decision-making problem under a Contextual Markov Decision Process (CMDP)(64). CMDP allows explicit modeling of longitudinal treatment-response data for heterogeneous patients post-stroke via context-dependent dynamics. We further extended the CMDP framework with realistic treatment constraints (e.g., the total dose and maximum bi-weekly dose). Finally, towards a concrete implementation of our rehabilitation system, we proposed a novel Posterior Sampling for Contextual RL (PSCRL) algorithm to balance exploration and exploitation in a CMDP while tackling the treatment constraints, which can be specified dynamically by the clinician-patient team at any time.

Importantly, the PSCRL algorithm is continuously learning (i.e., self-improving) at the patient and population levels by leveraging a growing database containing data from current and previous patients. We then presented a simulation study applying PSCRL to treat a highly variable population of 100 patients. The simulation utilized a previous hierarchical Bayesian state-space patient model of chronic stroke (29) expanded to include a context made of four covariates. Our results confirmed the multi-level learning capability of PSCRL as its recommended treatment plans became increasingly more effective in improving functional gains for each patient over time and across the patient population.

A strength of the Bayesian approach for patient modeling is that it provides a simple solution to missing data, which are treated as parameters to estimate. For instance, the model can be designed to incorporate covariates derived from neuroimaging to generate precise predictions. If, however, there is no brain scan available for a particular patient, the predictions may be (slightly) less accurate, but all the other data for this patient can still be used without any changes to the model. Thus, the Bayesian approach provides great flexibility for predictions with whichever data are available at any time for a specific patient. Further, hierarchical modeling, by sharing information across patients, improves prediction for new patients, especially early in the rehabilitation process, and therefore improves the efficacy of treatments.

### Limitations and future work

In future work, a large and variable rehabilitation dataset will be essential to train and validate the AI algorithm, notably by identifying the scheduling and individual factors that significantly affect outcomes. Hundreds of participants with large ranges of initial deficits will be needed. The database will contain baseline demographics, clinical, and lesion covariates, repeated clinical outcome measurements (i.e., every two weeks), and fine-grained practice data. Connected objects for rehabilitation (79), wearable sensors (80), or rehabilitation robots (41) can provide doses of treatment (in number of repetitions) for different tasks that can be used to update the model dynamics.

These data will be used to update the model, as the synthetic patients in the current work were represented by simple models while actual patients are much more complex, variable, and harder to model. Further extension of the patient model to acute and sub-acute phases post-stroke should include a time-dependent “plasticity state” to model the critical window of plasticity as well as the spontaneous recovery state, both modulated by covariates. In addition, in our current implementation, the algorithm made simplistic dose recommendations. Practically, the advantage of this approach is that only the actual dose in hours of treatment would need to be recorded and inputted into the model. However, in practice, treatment needs to be targeted to address the main limitations in UE function and preferences for specific activities for the patient to practice. Thus, an extension of our algorithm should at least provide recommendations for targeted training, such as for distal and proximal UE tasks.

The hierarchical Bayesian patient dynamics model used in our simulations was developed for chronic stroke UE functions based on a subjective patient-reported instrument, the MAL. Future models should consider widely used, objective, and validated functional clinical assessment scores, such as the ARAT, and account for the variability in recovery in impairment, function, and participation post-stroke. In a concurrent work, Cotton et al. (2024)(81) recently presented a related perspective to precision rehabilitation, advocating for the use of structural causal model for treatment optimization. The causal models would link the plastic process and neural structure underlying the rehabilitation process to detailed measurements of impairment of body structure and function to real functional activities of daily living and participation. Whereas the data requirement for such complex models would be very large, causal models, thanks to their transportability property, can be fit to a mixture of heterogeneous data.(81) Nonetheless, we note that MDP (and notably POMDP introduced below) encapsulates state-space models (such as (29)) and can be seen as simple structural causal models focusing on the interaction between a few variables at each timestep. Structural causal models can further augment MDPs, e.g., by decomposing the state and imposing structural causal assumptions among state components and the reward (82).

In our simulation, we assumed no measurement noise to accommodate the MDP model with fully observed states, i.e., *o*_*i,t*_, = *s*_*i,t*._ For real-world deployment, it could be beneficial to consider a stochastic observation function 𝕆(*o*_*t*|_*s*_*t*_) to reflect the measurement errors in the clinical outcomes (e.g. ARAT and MAL); this corresponds to a Partially Observable MDP (POMDP) where the patient state *s*_*i,t*_ (e.g., motor memory) is hidden from the clinician-AI team and partially revealed through the outcome *o*_*t*_. However, RL in a POMDP is known to be computationally intractable in general. In particular, a theoretically optimal policy for POMDP needs to be full-history dependent, i.e., it suggests treatments based on the entire patient history. In practice, when a single observed outcome *o*_*i,t*_ is insufficient for making a clinical decision, one may consider treatment policies that take as input a sequence of measurements from a few previous timesteps.

We finally note that POMDPs for precision rehabilitation are highly related to previous work on Dynamic Treatment Regime (DTR)(83). As reviewed in(84), constructing a DTR involves solving a decision problem that is mathematically equivalent to a POMDP, and indeed an optimal DTR is equivalent to an optimal (i.e., reward-maximizing) policy under a POMDP. Thus, we may interpret PSCRL as an algorithm that aims to *learn* the optimal DTR in a CMDP where the recovery dynamics are unknown and patient-dependent. Similar to PSCRL, a posterior sampling-based algorithm, PS-DTR, has been applied to the “causal RL” problem of learning an optimal dynamic treatment regime for an unknown structural causal model (85). Future work is needed to validate and compare these two algorithms in identifying the optimal rehabilitation plan in real life with high data efficiency and low regret, i.e., the least amount of trial and error. Note that these two RL algorithms are “online” in the sense that they continuously adjust the current treatment plan based on the patient data, and the updated plans are deployed to generate new observations. Online RL is suitable for precision rehabilitation as the interventions pose minimal risk; the clinician retains the autonomy for making treatment decisions and may reject the algorithm’s proposal at any time. Additional safety can be guaranteed by adjusting the constraints in our framework, in which case PSCRL can generate updated treatment plans accordingly.

## Conclusion

With self-improving capabilities, our AI-based system has the potential to play a key role in novel learning health systems in rehabilitation. As discussed above, stroke outcomes vary significantly based on factors such as the type, location, and severity. Making progress in precision medicine is crucial in stroke rehabilitation because it will enable the creation of personalized treatment plans tailored to each patient’s unique needs. Our collaborative AI system can transform stroke rehabilitation into a more adaptive and dynamic process, significantly improving patient outcomes and expanding the possibilities of personalized healthcare by providing a translatable framework for other clinical fields in which repeated treatments are needed to optimize outcomes.

## Data Availability

All data produced in the present work are contained in the manuscript

## Declarations

### Ethics approval and consent to participate

Not applicable

### Consent for publication

Not applicable

### Availability of data and materials

Not applicable

## Authors’ contributions

DY and NS conceived the work and contributed to the design of the work, data interpretation, and drafting of the manuscript. DY developed and implemented the PSCRL algorithm, performed the analyses, and ran the simulations. HL contributed to the algorithm development and revision of the manuscript. CW conceived the work, contributed to the design of the work and revision of the manuscript. All authors read and approved the final manuscript.

## Acknowledgments

We thank David Reinkensmeyer, Emily Rosario, and Sook-Lei Liew for fruitful discussions about our framework and Rahul Jain for discussion about the PSCRL algorithm.

## Footnotes

a Other neural factors include such as transcallosal tract integrity, ipsilesional motor cortex activity, and connectivity between motor and premotor cortex (41, 45, 53-56). However, we note that identifying these factors requires TMS, EEG, or research-grade MRIs, which are often unavailable in routine care.

b We defer the technical details of PSCRL and a review of the related theoretical literature to Supplementary Material A.

c We reserve *s*_*i,t*_ and 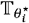 for the state and transition function of an augmented MDP used by the RL agent for treatment planning. This is because the agent’s “perceived” state *s*_*i,t*_ may carry more information (e.g., the remaining budget).

