## Supplementary Material for "Towards AI-based Precision Rehabilitation via Contextual Model-based Reinforcement Learning"

### A. CMDP Technical details

We formulate precision rehabilitation as a decision-making problem of optimizing rehabilitation treatments. The formalism follows a contextual Markov decision process (CMDP) in the RL literature. CMDP was introduced )(1, 2) as an extension to the Markov decision process (MDP). An MDP can be thought of as a controlled Markov chain (a simple state-space model) where the evolution of a system’s (in our case, patient’s) state depends only on its current state and agent’s (or clinician’s) action.

**Notations.** We first reiterate necessary notations as defined in the main text. For any positive integer $m$, we define $\left[ m \right]=\{1,2,\ldots,n\}$. For any two positive integers $n\leq m$, we use the shorthand $n:m=\{n,n+1,\ldots,m\}$. We denote an ordered collection of values (or random variables) by applying this notation in subscript. For instance, we write $s_{i, 1:H}=\left( s_{i,1},s_{i,2},\ldots,s_{i,H} \right)$ to denote a sequence of states for patient $i\mathbb{\in N}$until timestep $H\mathbb{\in N}$. For $d$ arbitrary real numbers $a_{1},a_{2},\ldots,a_{d}$, we also use $a_{1:d}=\left( a_{1},a_{2},\ldots,a_{d} \right)$ to denote a $d$-dimensional vector, i.e., $a_{1:d}\in\mathbb{R}^{d}$.

**Markov decision process (MDP).** Formally, a finite-horizon MDP is defined by a tuple $\left( \mathcal{S,A,}\mathbb{T,}H,R,d_{1} \right)$:

1. $\mathcal{S}$ is a *state space*, a set of states characterizing the status of the *environment* (i.e., the patient currently taking treatments) at a given timestep.
2. $\mathcal{A}$ is an *action space*, a set of actions (or treatments) available to the *agent* (i.e., the clinician).
3. $\mathbb{T}$ is a *transition (probability) function* such that $\mathbb{T}\left( s^{'} | s,a \right)$ gives the probability that starting from state $s\in\mathcal{S}$, the environment transforms into state $s^{'}\in\mathcal{S}$ as a response to action $a\in\mathcal{A}$ performed by the agent (after one timestep).
4. $H$ is the *horizon*, the fixed number of stages or timesteps over which the interaction occurs.
5. $R:\mathcal{S\times A\times S}\mathbb{\to R}$ is a *reward function* where $R(s,a,s^{'})\mathbb{\in R}$ is the reward received by the agent after reaching next state $s'\in\mathcal{S}$ by performing action $a\in\mathcal{A}$ at state $s\in\mathcal{S}$.
6. $d_{1}\in\Delta\left( \mathcal{S} \right)$ is an *initial state distribution* for the environment (or the patient).

Notice that components of an MDP may vary in time (e.g., by adding a subscript $t$). In the simulation below, a time-varying reward function $R_{t}$, where $R_{t}(s_{t},a_{t},s_{t+1})$ signifies the instantaneous reward given to the agent at time $t$ after reaching state $s_{t+1}$ by performing action $a_{t}$ at state $s_{t}$.

**Contextual MDP (CMDP).** To introduce context-dependent patient-level dynamics, we consider a CMDP defined by a tuple $\left( \mathcal{S,A,}\mathbb{T}_{\theta},H,R,d_{1}\mathcal{,C,}\Theta, \phi\right)$ with the following additional component:

7. $\mathcal{C}$ is a *context space*, a set of contexts that may aid the agent in identifying the dynamics in the current episode.
8. $\Theta$ is a *parameter space*, a set of values for the parameter that fully characterizes the transition probabilities associated with each patient.
9. $\phi$ is either a) a conditional distribution of parameter $\theta\in\Theta$ given context $c\in\mathcal{C}$, or b) a function that deterministically maps a context $c\in\mathcal{C}$ into a parameter $\phi\left( c \right)=\theta_{c}\in\Theta$.

Moreover, CMDP involves a parameterized transition probability function:

$3^{\star}$. $\mathbb{T}_{\theta}\left( s' |s,a \right)$ gives the probability of reaching $s^{'}\in\mathcal{S}$ from $s\in\mathcal{S}$ by performing $a\in\mathcal{A}$ for the patient with parameter $\theta\in\Theta$.

It is not necessary to impose a distributional assumption on the arriving patient sequence, represented by a sequence of observable contexts and hidden parameters. In particular, the patient sequence may be *adversarial* in the sense that an expert clinician would (and perhaps should) treat harder cases. In this work, however, for simplicity we considered a *stochastic* patient sequence with a population distribution $\nu_{c}$ of contexts.

The goal of an agent in CMDP is to find an optimal behavior policy. For a finite horizon problem, we consider a non-stationary policy, which is defined by a time-indexed function $\pi=\left( \pi_{1},\ldots,\pi_{H} \right)$ mapping states to actions, where $\pi_{t}\left( s \right)\mathcal{\in A}$ gives the action to be performed if the patient’s state is $s\in\mathcal{S}$ at timestep $t\in[H]$.

**Budget constraints.** We further introduce budget constraints to the precision rehabilitation problem. Given an integer-valued total budget  $\bar{B}$, a budgeted policy is denoted by $\pi^{\bar{B}}=\left( \pi_{1}^{\bar{B}},\ldots,\pi_{H}^{\bar{B}} \right)$ where $\pi_{t}^{\bar{B}}\left( s,b \right)\in\mathcal{A}$ is the action to be performed if, at time $t$, the patient’s state is $s\in\mathcal{S}$ and the remaining budget is $b\in\left[ 0:\bar{B} \right]=\{0,1,\ldots,\bar{B}\}$. We call $\left( s,b \right)\in\mathcal{S}\times\left[ 0:\bar{B} \right]$ an *extended state* for the budgeted MDP as the patient’s state $s$ and the remaining budget $b$ are both necessary information for selecting treatments.

**Interaction protocol.** The CMDP model assumes that a sequence of patients arrives independently. We treat each patient $i=1,2,\ldots,$ for a fixed treatment window of $H$ stages and then move to patient $i+1$. The treatment window for a patient is called an *episode* of the CMDP. In episode $i$, for each timestep $t=1, 2, \ldots, H$, we observe the current state $s_{i,h}$ of patient $i$ and then choose a treatment $a_{i,t}$ for this patient based on the available data, the reward function $R$, and a learning strategy. The next state $s_{i,h+1}$ is assumed to follow the dynamics model $\mathbb{T}_{\theta_{i}}$, i.e., $s_{i,t+1}\sim\mathbb{T}_{\theta_{i}}\left( \cdot| s_{i,t},a_{i,t} \right)$, where $\theta_{i}\in\Theta$ is a hidden parameter that characterizes the (conditional) distribution of $s_{i,t+1}$ given $s_{i,t}$ and $a_{i,t}$. After performing $H$ actions on patient $i$, we observe the terminal state $s_{i,H+1}$ and reward $r_{i, H}=R_{H}\left( s_{i,H},a_{i, H},s_{i,H+1} \right)$ and episode $i$ terminates. The data collected throughout episode $i$ is called a *trajectory* $\tau_{i}=\left( s_{i,1},a_{i,1},s_{i,2},\ldots,s_{i,H},a_{i,H},s_{i,H+1} \right)$.

**Connection to HBDM.** We previously proposed a hierarchical Bayesian dynamics model (HBDM) as a predictive model of the time course of Motor Activity Log (MAL) based on the DOSE and EXCITE trials (3). A CMDP may be deemed as a control (or RL) theoretic extension of the corresponding dynamics model. Specifically, the transition function in CMDP is mathematically equivalent to a HBDM with an additional context-dependent dynamic. As introduced in section [Algorithm], our model-based RL algorithm additionally incorporates Bayesian inference of the learning of unknown, context-dependent dynamics for each patient.

### B. Detailed Simulation specification

We implemented a CMDP simulator for the univariate dose optimization problem with following specifications:

**States.** We used $\mathcal{S=}\left[ 0, 5 \right] \times\left[ 0:60 \right]$ such that for the state $s_{i,t}=\left( o_{i,t},b_{i,t} \right)\mathcal{\in S}$ of patient $i$ at timestep $t$, the first coordinate $o_{i,t}$ corresponds to the patient’s current MAL measurement, with a minimum score of 0 and maximum score of 5, and the second coordinate $b_{i,t}\in[0:60]$ is the remaining rehabilitation budget (i.e., an upper limit on the dose for the current treatment).

**Treatments.** As the rehabilitation dose is assigned at integer amounts, we set $\mathcal{A=}\left[ 0:20 \right]$ and let $a_{i,t}\in\mathcal{A}$ be the rehabilitation dose (i.e., hours of therapy) for patient $i$ at time $t$. (We enforced the budget constraint in the reward function.)

**Contexts.** For each synthetic patient $i=1,2,\ldots,N$, we generated a four-dimensional context vector $c_{i}=c_{i, 1:4}\in\mathbb{R}^{4}$ (i.e., four covariates) according to a population context distribution:

$$c_{i,1}\sim\text{Normal}\left( \mu=24,\sigma^{2}=16 \right)$$

$$c_{i,2}\sim\text{Uniform}\left( 2, 60 \right)$$

$$c_{i,3}\sim\text{Bernoulli}\left( 0.3 \right)$$

$$c_{i,4}\sim\text{Bernoulli}\left( 0.5 \right)$$

where the components within each $c_{i}$ are mutually independent, and across patients, $c_{i}$’s are independent and identically distributed.

**Patient-level dynamics.** As in the main text, we use superscript “$\star$” to denote a ground-truth quantity unknown to the agent. As in Schweighofer et al. 2023, we parameterized patient $i$’s dynamic by $\theta_{i}^{\star}=\left( \alpha_{i}^{\star},\beta_{i}^{\star},\gamma_{i}^{\star} \right)$, where $0<\alpha_{i}^{\star}<1$, $\beta_{i}^{\star}\geq0$, and $\gamma_{i}^{\star}\geq0$ are retention, learning, and self-training rates respectively. We made the conditional independence assumption that $\theta_{i}^{\star}$ only depends on the context $c_{i}$ up to some random effect. We imposed a deterministic relationship from $c_{i}$ to the retention and self-training rates ($\alpha_{i}^{\star}$ and $\gamma_{i}^{\star}$), while setting a stochastic relationship from $c_{i}$ to the learning rate ($\beta_{i}^{\star}$) to allow further variability in treatment effects. Given $c_{i}\in\mathbb{R}^{4}$, the (ground-truth) parameter $\theta_{i}^{\star}=\left( \alpha_{i}^{\star},\beta_{i}^{\star},\gamma_{i}^{\star} \right)$ for patient $i$ was generated according to the following (conditional) distribution:

$$\theta_{i}^{\star}\sim\text{Normal}_{\Theta}\left( f\left( c_{i} \right),\Sigma_{\theta}^{\star} \right)$$

$$f\left( c_{i} \right)=\left[ \begin{matrix} {w_{\alpha}^{\star}}^{⊺} \\ {w_{\beta}^{\star}}^{⊺} \\ {w_{\gamma}^{\star}}^{⊺} \end{matrix} \right]\left[ \begin{aligned} 1 \\ c_{i} \end{aligned} \right]=\left[ \begin{matrix} .8 & .004 & 0 & -.1 & 0 \\ .1 & 0 & .01 & 0 & .1 \\ .3 & -.01 & .01 & 0 & 0 \end{matrix} \right]\left[ \begin{aligned} 1 \\ c_{i} \end{aligned} \right]$$

$$\Sigma_{\theta}^{\star}=\text{diag}\left( 0,.04,0 \right)$$

where $\text{Normal}_{\Theta}$ denotes a *truncated (multivariate) normal distribution* with respect to the boundary of the parameter space

$$\Theta=\{\left( \alpha,\beta,\gamma\right)\in\mathbb{R}^{3}:0<\alpha<1, \beta\geq0, \gamma\geq0\};$$

$f:\mathbb{R}^{4}\to\Theta$ is a linear function with ground-truth parameters $w_{\alpha}^{\star}=w_{\alpha,0:4}^{\star}$, $w_{\beta}^{\star}=w_{\beta,0:4}^{\star}$, and $w_{\gamma,0:4}^{\star}$, where $w_{\alpha,1:4}^{\star},w_{\beta,1:4}^{\star},w_{\gamma,1:4}^{\star}$ correspond to fixed effects for each covariate, and $w_{\alpha,0}^{\star},w_{\beta,0}^{\star},w_{\gamma,0}^{\star}$ correspond to fixed intercepts; and covariance matrix $\Sigma_{\theta}^{\star}$ characterizes the scale of random effects on the dynamics that are not explained by the context.

**Patient-level dynamics without random effect.** For comparison, we also constructed a simulator with a deterministic context-to-dynamics relationship by removing the random effect, i.e., $\theta_{i}^{\star}=f\left( c_{i} \right)$, where that $c_{i}$ is appropriately generated so $\theta_{i}^{\star}$ is a feasible parameter. This corresponds to the scenario where the recovery dynamic can be identified directly as the patient-specific context is informative enough. Although a stochastic context-to-dynamics relationship is more realistic, we note that with sufficient context data and a deep neural network, it is still a common practice to simply train deterministic predictors and plan accordingly. Even if the underlying context-to-dynamics relationship is deterministic, learning such a relationship is still a difficult task since the time course of patient’s states is stochastic in nature and the recovery dynamics ($\theta_{i}$’s) are not directly observed by the agent.

**Initial state.** We considered patients arriving with random initial clinical states. For patient $i$, the initial state $s_{i,1}=\left( o_{i,1},b_{i,1} \right)$ contains an initial MAL outcome $o_{i,1}$ randomly generated from $\text{Normal}_{\left[ 0,5 \right]}\left( 2, 0.2 \right)$, and for ease of comparison between treatment policies, a fixed initial (total) budget $b_{i,1}=60$.

**Trajectories.** Given the initial MAL $o_{i,1}$, the time course of MAL $\left( o_{i,1:H+1} \right)$ was generated with a non-linear dynamical system specified by:

$$x_{i,t+1}\sim\text{Normal}\left( \mu=\alpha_{i}^{\star}x_{i,t}+\beta_{i}^{\star}a_{i,t}+\gamma_{i}^{\star}o_{i,t},\sigma^{2}=0.25 \right)$$

$$o_{i,t}=5\cdot\text{Sigmoid}\left( 0.2x_{i,t}-3 \right)$$

for each patient $i=1,2,\ldots,N$ and each timestep $t=1,2,\ldots,H$. The doses ($a_{i,t}'s)$ are decision variables given as input to the simulator at each round of treatment. The budget component $b_{i,t}$ of a patient state transitions deterministically such that $b_{i,t+1}=b_{i,t}-a_{i,t}$. To emulate the 6-month treatment window the treatment horizon was set to $H=12$, where each timestep corresponds to a 2-week interval. The sample size was set to $N=100$, but we note that $N$ could be arbitrarily large since the simulation may run indefinitely long.

### C. Additional results: covariate parameter estimation

Fig. S1 shows the evolution of the posterior distributions, as a function of the number of participants, of the context-to-dynamics parameters estimated by PSCRL. We plotted the posterior distribution (with mean and 95% credible intervals) for the weight matrix $W$ of the linear context-to-parameter relationship (see Tables 1 and 2) once per patient (at the start of each treatment episode, i.e., $t=1$) although posterior updates were performed at every timestep. For most parameters, the posterior distribution concentrated around the ground-truth after treating around 20 patients. The only exceptions were that the posterior for weights $w_{\gamma,3}$and $w_{\gamma,4}$ did not shrink with more data (where the ground truth values were $w_{\gamma,3}^{\star}=0$and $w_{\gamma,4}^{\star}=0$). Overall, the history of the posterior distribution showed that PSCRL achieved learning on the population-level and mostly recovered the context-to-parameter relationship.

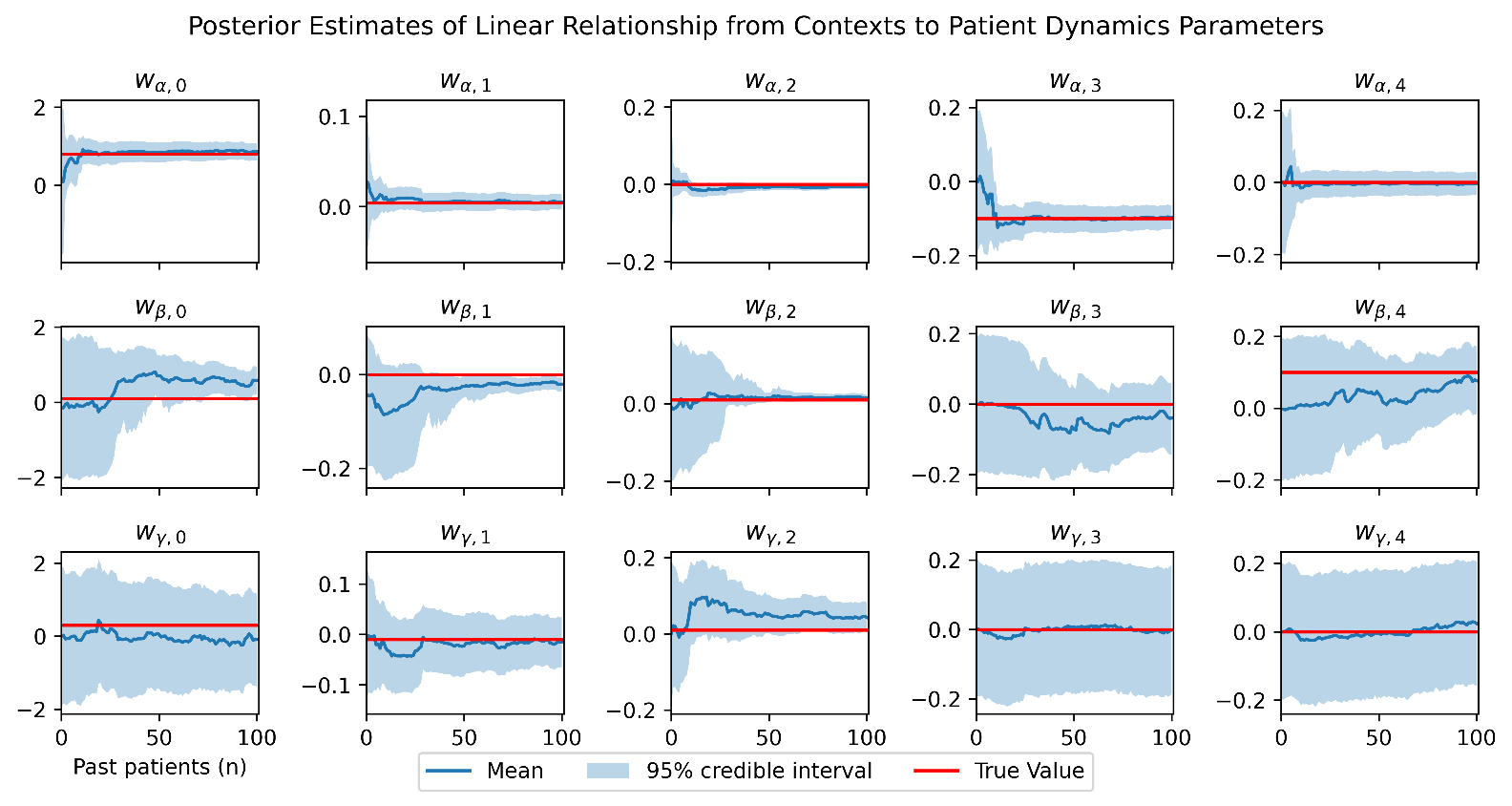

Fig. S1. Learning the fixed effects (and intercepts) from contexts to latent dynamics parameters.

1.	Hallak A, Di Castro D, Mannor S. Contextual markov decision processes2015.

2.	Modi A, Jiang N, Singh S, Tewari A. Markov decision processes with continuous side information. arXiv preprint arXiv:171105726. 2017.

3.	Schweighofer N, Ye D, Luo H, D'Argenio DZ, Winstein C. Long-term forecasting of a motor outcome following rehabilitation in chronic stroke via a hierarchical bayesian dynamic model. J Neuroeng Rehabil. 2023;20(1):83.
